# Testosterone replacement therapy ameliorates spatial cognitive function in age-related hypogonadism (LOH) patients: analysis with a virtual three-dimensional maze

**DOI:** 10.1101/2025.01.27.25321222

**Authors:** Suguru Kawato, Hideo Mukai, Yusuke Kai, Masaki Kimura, Mari Ogiue-Ikeda, Mika Soma, Minoru Saito, Shigeo Horie

## Abstract

When comparing the behavioral levels of men and women using a maze navigation game created on a PC to investigate spatial cognitive function, many reports indicated that men are significantly faster than women at finding the exit of the maze.

Analysis with functional MRI neuroimaging indicates that neural activation at multiple sites were observed during a maze navigation task. Significantly different regions were identified in men and women, with men showing significantly increased activity in the hippocampus. In contrast, women showed increased activity in the Brodmann area of the prefrontal cortex. When differences in the activated regions were taken between men and women, the hippocampal activity showed the largest difference.

Since the hippocampal CA1 regions are known to be essentially responsible to spatial navigation ability, the reported difference in spatial ability between men and women could be due to higher testosterone (T) levels in mem than in women, because the application of T can increase hippocampal CA1 neural synapses as observed in rats, for example.

In the present study, we developed the original 3D maze navigation test on a PC to accurately analyze spatial cognitive function, and this maze was used to analyze differences in spatial cognitive function between young male and female volunteers. The results showed that males took a distinctly shorter time and path length to escape the maze than females. We applied this maze test to analyze the possible improvement effects of spatial cognitive function by T-replacement therapy (TRT) on male patients with late onset hypogonadism (LOH). LOH patients have significantly low levels of T as well as high Aging males’ symptoms scale (AMS) scores when compared with healthy men. Upon TRT for 6 weeks, we revealed that the spatial cognitive ability of LOH patients was significantly improved along the increase of T levels and improvement of AMS scores.

The observed improvement of spatial cognitive ability of LOH patients could be interpreted as effects of elevation of T levels in the hippocampal CA1 region where androgen receptor (AR) is localized.

## Introduction

Continuous visuospatial navigation in familiar and unfamiliar environments is a requirement of daily life.

Visuospatial navigation is one of the few cognitive functions for which a reliable gender-specific performance difference is well known (Astur et al 1998). For example, men were significantly faster than women at finding the way out of maze.

Navigation through mazes can be combined with functional neuroimaging for men and women. From the fMRI analysis, gender differences in brain neural activation was computed with maze navigation tasks for male and female subjects (Grön et al 2000). Although there was great overlap between men and women in brain regions that were significantly activated, there were brain regions that were activated exclusively by the male and female groups. From the comparison of the neural activity, men showed significantly increased activity in the hippocampus proper, however, women showed increased activity of the prefrontal cortex, in contrast with men. (Grön et al 2000)

In the hippocampus, the glutamatergic neurons in the CA1 region are responsible for spatial cognition (Tonegawa et al 1996). This is evidenced by the fact that spatial cognition is selectively inhibited in mice in which N-methyl-D-aspartate (NMDA) receptors of glutamatergic neurons in the CA1 region of the hippocampus are specifically knockout (Tsien et al 1996) (McHugh et al 1996).

Previous studies of sex hormones in rats have shown that the level of testosterone (T) in the hippocampus is much higher in males (approximately 17 nM) than in females (approxi. 0.05 nM) (Hojo et al 2009) (Ooishi et al 2012) (Kato et al 2013). T activates neurons upon binding to androgen receptor (AR). AR is significantly expressed in glutamatergic neurons of the hippocampal CA1 region (Tabori et al 2005) (Hatanaka et al 2015). In addition, T level is much higher in male hippocampus than female hippocampus (Kato et al 2013). Circulating T levels are qualitatively proportional to hippocampal T levels (Kato et al 2013) (Hojo et al 2014) (Hojo & Kawato 2018). In view of these situations, we hypothesize that “higher T levels in males and lower T levels in females may have a significant effect on the difference in spatial cognitive function between men and women”.

In Section 1, based on this hypothesis, we created a virtual maze escape game on a PC in order to find the spatial cognitive ability that may depend on T levels.

In Section 2, we applied this maze whether this maze is able to clearly determine differences in spatial cognitive ability between young men and women. We obtained the results that young men were significantly faster than young women in escaping from the maze.

In Section 3, we applied this maze navigation test to male patients with late onset hypogonadism (LOH) in order to examine whether T-replacement therapy (TRT) is effective to improve spatial cognitive ability by increase of T levels. It should be noted that because LOH patients have low T levels, TRT can elevate T levels of LOH patients, and thereby can improve Aging males’ symptoms scale (AMS) scores (Heinemann et al 2003) (Lunenfeld et al 2013) (Ide et al 2023). However, there had not been any challenges to investigate how TRT could improve spatial cognitive ability of LOH patients.

## Section 1: Creation of 3D maze for spatial maze navigation test

### 1.1 Technical specification of maze

A 3D virtual maze program was created using the 3D modeling software Unity 5.6.1 (Unity Technologies). C# was used as the programming language.

The PC environment for the maze program and operation is as follows

S: Windows 10 or 11, 64bit
CPU: Intel Core i7-6700HQ 2.60GHz
RAM: 16.0GB
GPU: Intel HD Graphics 530
Monitor: 15.6 in wide

The size of the maze is 6 squares (vertical) x 6 squares (horizontal) for a total of 36 squares (Figure 1.1, 1.2).

**Figure 1.1.**
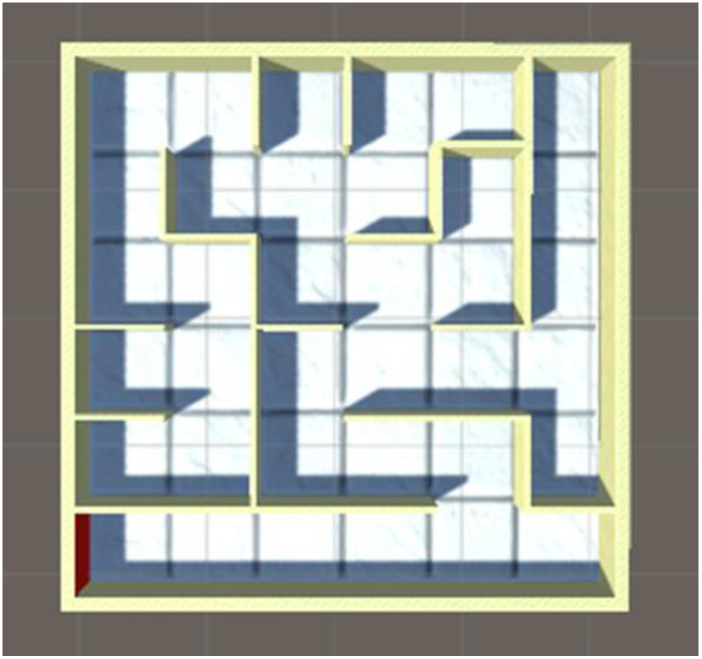
Aerial view of the maze. The maze is 6×6 squares in size and can be presented in a choice of 15 different ways.

**Figure 1.2.**
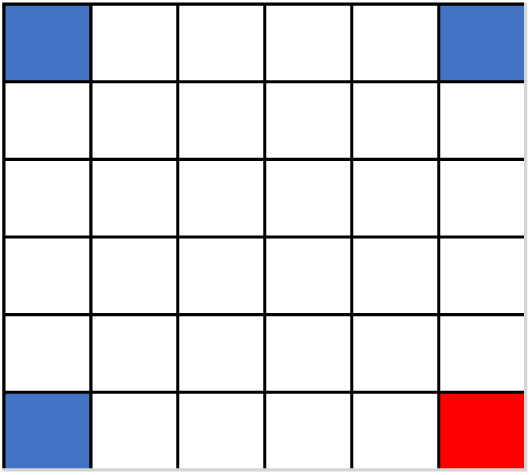
How the start (blue) and goal (red) points are selected. If the start point is in a red square, the goal point is randomly selected from the blue squares. Note that subjects cannot see the goal that is hidden from the start by a wall.

Fifteen sets of maze (Maze Number 1 – 15) are prepared. One of the maze can be selected by choosing the group number (1, 2 or 3) on the title screen (see below), and then automatically one Maze will be randomly selected. Note that one group consists of 5 mazes depending on goal positions.

#### Start and goal points

The 15 mazes are divided into three groups: one with a goal in square (6, 6), one with a goal in square (1, 6), and one with a goal in square (6, 1). Subject can choose which maze with a goal position you want to generate by entering the maze number on the title screen (see below).

The start point is randomly selected from the four corners of the maze except for the goal. For example, in Figure 1.2, the goal is located at square (6, 6) (red square), so the starting point is chosen from the squares (1, 1), (1, 6) and (6, 1) (blue square).

#### Operation of maze task

##### Start, top title screen

When the maze navigation program is started, the top title screen appears. On the title screen, the subject enters his/her name and maze number in order to select the type of maze. Two types of mazes are available: maze without landmarks or maze with three landmarks.

Before starting the maze test at the first time, “Demo Video” should be selected on the title screen in order to get an overview of the present navigation maze task.

When ‘walking’ through the maze, subjects have a first-person view. During the maze test, the arrow keys on the keyboard can be used to move forward with the up arrow key, backward with the down arrow key, and turn right and left with the right and left arrow keys, respectively. Turning was possible within the full range of 360 degree. [↑] forward, [↓] backward, [→] right turn, [←] left turn.

##### Selection of the time limit screen

Once the time limit is selected, the maze test begins. When the time limit is exceeded, the word “End” is displayed.

During the maze test, a subject proceeds in the first-person perspective and searches for the correct path.

##### Goal screen

When a subject reaches the goal (the red wall) (Figure 1.6), a subject should presses the return key to finish the task. Then the time taken to reach the goal and the distance traveled during the maze task are recorded. At the same time, the screen will be back to the title screen (see Fig 1.3).

**Figure 1.3.**
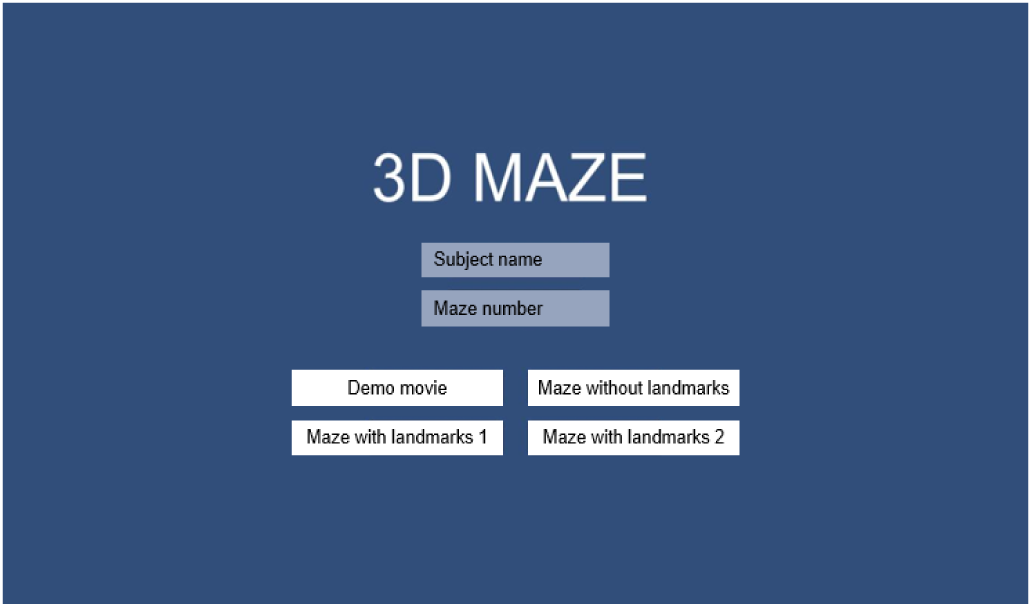
Top title screen. Press Return to next Selection screen.

**Figure 1.4.**
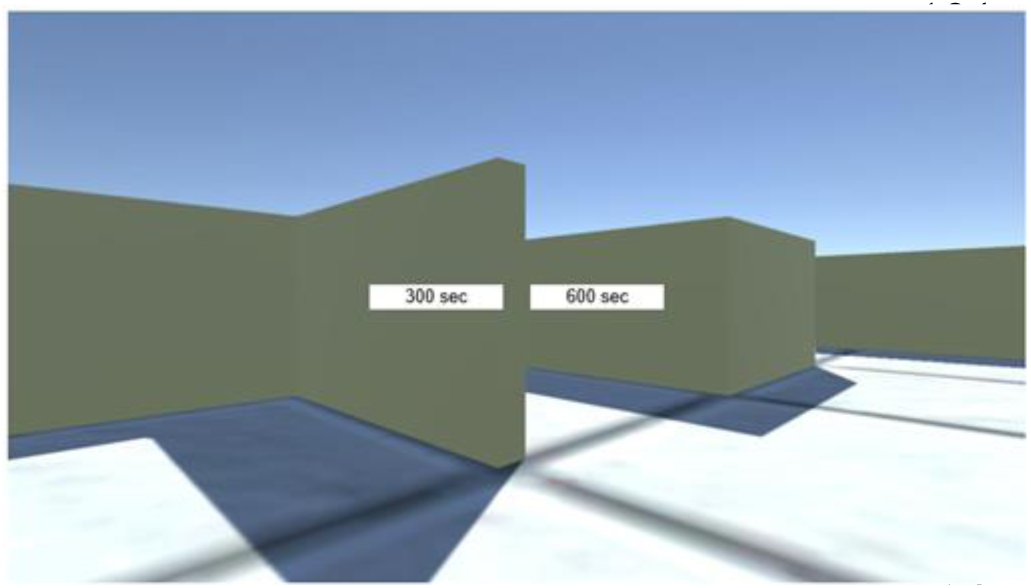
Time limit selection screen. There are two time limits (300 and 600 seconds) that can be chosen. Normally 300 sec is chosen.

**Figure 1.5.**
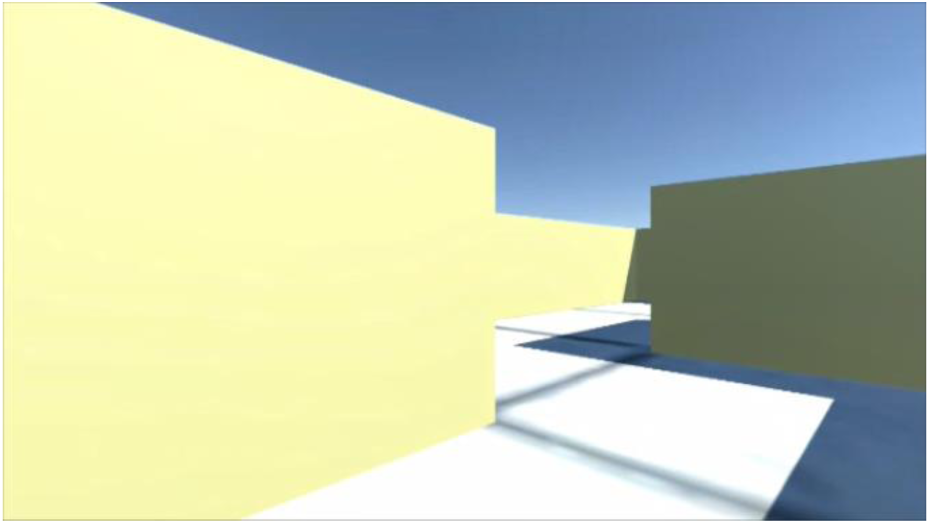
Typical view inside the maze. During the control condition, the screen is frozen at the last position of the active condition.

**Figure 1.6.**
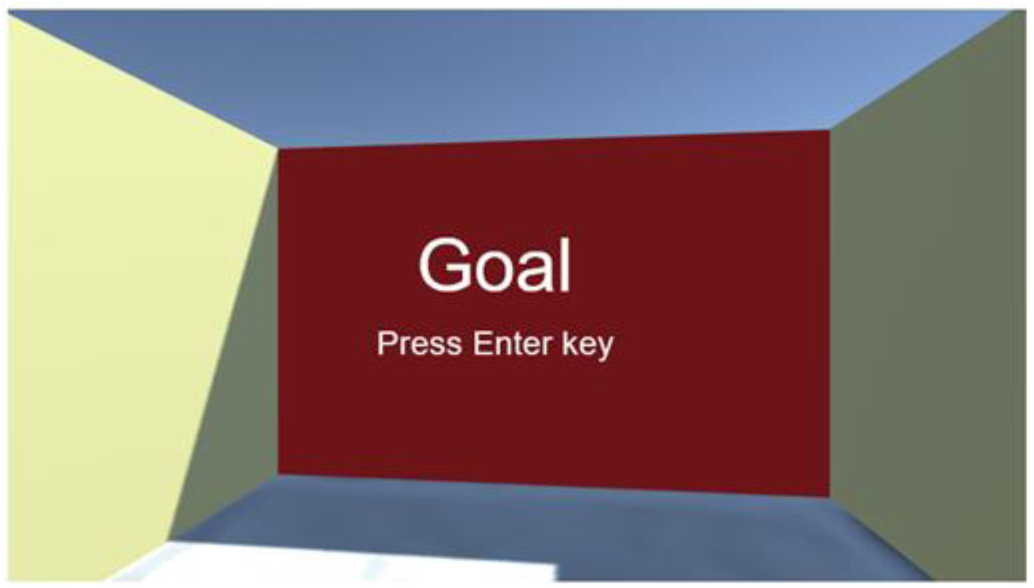
The “Goal” is displayed on the goal wall. Pressing the Enter key to finish.

**Figure 1.7.**
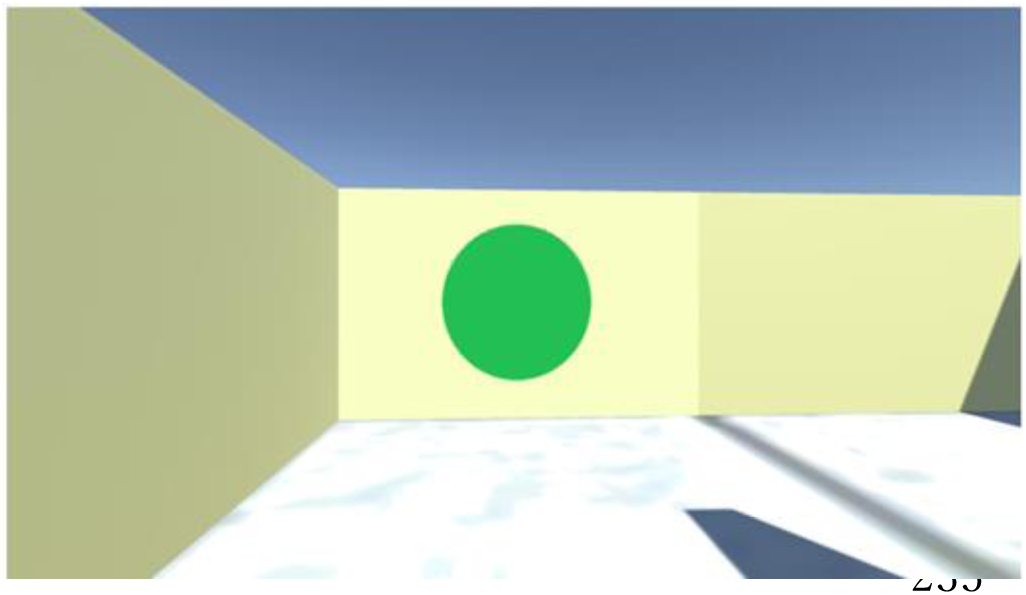
Example of landmarks in the maze. Three types of landmarks(circle, star, and triangle) are used. Only three walls have landmarks and each has different types of landmarks.

Landmarks in each maze are placed at the simple turned in the load to give a hint of location. Landmarks are not placed at intersection of the convey (that is, decision points).

### 1.2 Assessment of Spatial Cognitive Ability

#### Escape time

As a measure of spatial cognitive ability, the time taken to solve a maze test is measured. This is called escape time.

The escape time is the number of seconds from the start of the maze test (immediately after the time limit is selected) to the end of the maze test (when the subject reaches the goal or the time limit is reached). The escape time is output to the total score.csv file in the result folder directly under the C drive after the maze test is completed.

#### Path Distance

As a measure of spatial cognitive ability other than escape time, the distance traveled during the maze test is recorded. This is called the path distance.

The path distance, as well as the escape time, is output to the total score.csv file located in the results folder directly under the C drive after the maze test is completed. The Euclidean distance for each second during the maze test is taken, and the sum of these Euclidean distances from start to goal is defined as the path distance (transit distance).

## Section 2: Analysis of differences in spatial cognitive ability between young men and women using a virtual maze navigation task

### Materials and Methods

#### Subjects

The subjects were 40 healthy students (20 men and 20 women) affiliated with Meiji University. The ages of the men and women subjects were 21.6 ± 1.5 and 20.6 ± 1.2 years, respectively. The experiment was notified to and approved by the Ethics Committee on Safety and Human Subjects for Experimental Research on Genetic Recombination, School of Science and Engineering, Meiji University (Approval No.: RIKEN 14-520). Subjects were informed about the research in person in advance and their consent was obtained by signing a research participation consent form.

#### Experimental Procedures

The experiment will be conducted according to the following procedures.

1. After explaining the outline of the experiment, show a demonstration video of the maze test.
2. Ask the subjects to perform the maze escape task without landmarks.
3. Ask the subjects to perform the maze escape task with three landmarks.
4. Repeat steps 2 and 3 four times with an interval of one week.

When performing the maze test, maze number 1 should be entered in the first time, maze number 2 in the second time, maze number 3 in the third time, and maze number 4 in the fourth time, to avoid mazes with identical goal positions.

### Results

Weekly comparison of maze navigation task, using maze without/with three landmarks.

In the maze navigation task, young men significantly outscored young women in the maze without landmarks, however, this difference became smaller by the presence of landmarks (Figure 2.1).

**Figure 2.1.**
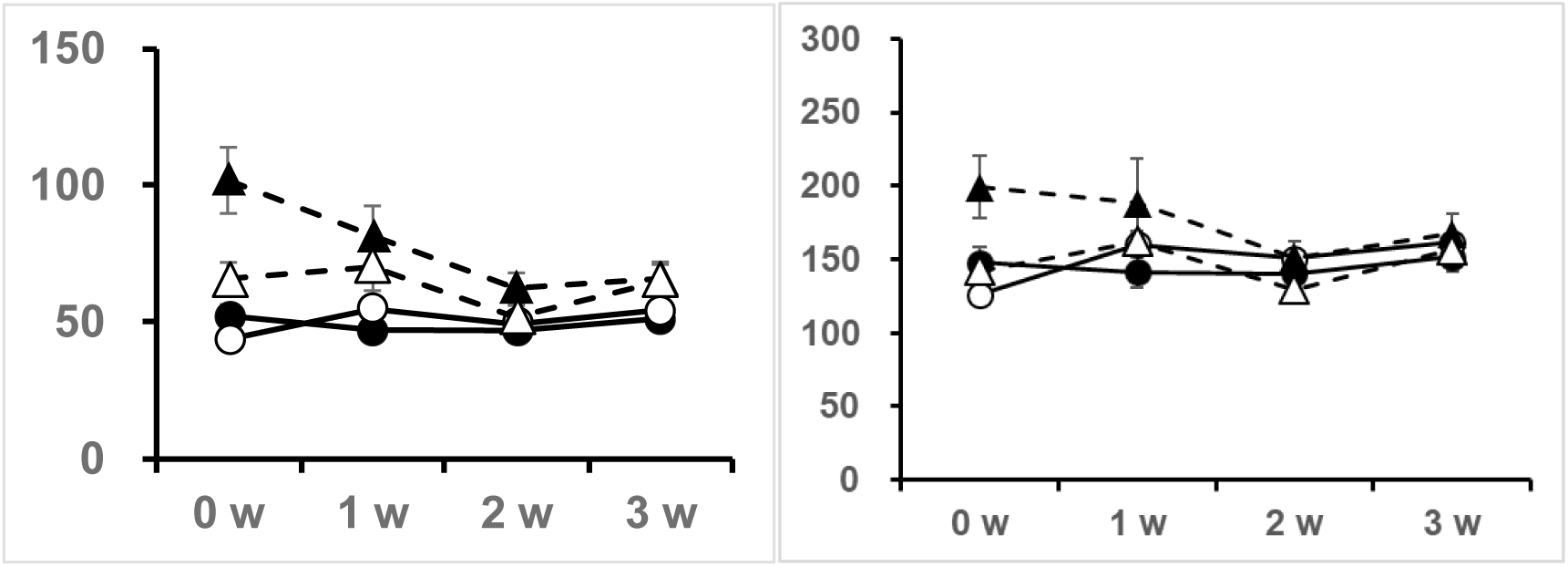
Weekly scores of escape time (Left) and path distance (Right) for young men and women in maze tests. Vertical axis is escape time (second) and path distance (arbitrary unit), respectively. (●)Young men without landmarks, (○) Young Men with landmarks, (▲) Young women without landmarks, (△) Young women with landmarks.

The escape time of young men are shorter than that of young women in both mazes without and with landmarks. However, the presence of landmarks made the difference smaller.

In terms of path distance, men showed significantly shorter path distance than women in the maze without landmarks, however, by the presence of landmarks women showed the same path distance as men (Figure 2.1). Differences between men and women are large in the absence of landmarks. Significant decreasing trends in escape time and path distance are observed only for women in the maze without landmarks.

Table 1.1 and Table 1.2 represent statistical analyses that showed a significance (p < 0.05) by comparison between gender (men or women), the presence and absence of landmarks and weeks.

**Table 1.1.** A two-way analysis of variance test revealed an interaction between gender (men or women) and presence/absence of landmarks for escape time and path distance, and an interaction between gender and week for escape time.

|  | Escape Time |  |  | Path Distance |  |  |
| --- | --- | --- | --- | --- | --- | --- |
| | *F* | *p* | $\eta^2$ | *F* | *p* | $\eta^2$ |
| Gender vs Presence/Absence of Landmarks | 8.31 | 0.0064** | 0.0176 | 6.59 | 0.0143* | 0.0147 |
| Gender vs Weeks | 4.50 | 0.0051* | 0.0344 | 1.41 | 0.244 | 0.0118 |
| Gender vs Presence/Absence of Landmarks vs Week | 0.789 | 0.503 | 0.00750 | 0.275 | 0.843 | 0.00270 |

\* $p < 0.05$ \*\* $p < 0.01$

**Table 1.2.** Simple main effect tests are conducted on escape time and path distance. In the analysis of gender and presence/absence of landmarks, effects are shown of gender in maze without landmarks, gender in maze with landmarks, and presence/absence of landmarks in women in escape time.

|  | Escape Time |  |  | Path Distance |  |  |
| --- | --- | --- | --- | --- | --- | --- |
| | *F* | *p* | $\eta^2$ | *F* | *p* | $\eta^2$ |
| Effect of Gender in Maze without Landmarks | 21.5 | < 1.00x10<sup>-4</sup>*** | 0.160 | 3.87 | 0.0564 | 0.0331 |
| Effect of Gender in Maze with Landmarks | 7.18 | 0.0108 * | 0.0598 | 0.252 | 0.618 | 0.00170 |
| Effect of Presence/Absence of Landmarks in Male | 0.0363 | 0.851 | 2.00x10<sup>-4</sup> | 0.0863 | 0.772 | 4.00x10<sup>-4</sup> |
| Effect of Presence/Absence of Landmarks in Female | 11.0 | 0.00370** | 0.0417 | 9.07 | 0.00720* | 0.0391 |
| Effect of Gender in 0w | 19.3 | 1.00x10<sup>-4</sup>*** | 0.224 |  |  |  |
| Effect of Gender in 1w | 11.5 | 0.00160** | 0.131 |  |  |  |
| Effect of Gender in 2w | 4.23 | 0.0467 * | 0.0629 |  |  |  |
| Effect of Gender in 3w | 1.85 | 0.182 | 0.0287 |  |  |  |
| Effect of Weeks in Male | 1.99 | 0.126 | 0.0268 |  |  |  |
| Effect of Weeks in Female | 4.86 | 0.00440** | 0.0741 |  |  |  |

\* $p < 0.05$ \*\* $p < 0.01$ \*\*\* $p < 0.001$

#### Weekly comparison in order to check learning effects on the maze test

During repeating the maze test, possible learning effects (with familiarity factors) might improve the skill to escape the maze. If the subjects become accustomed to the maze, their scores could be improved. We observed the decrease in the escape time and the path distance by repeating women’s maze tests without landmarks at different weeks (0th, 1st, 2nd, 3rd week trial) (see Figure 2.1). However, in women’s maze tests with landmarks, these improvement effects by repetitive training were not observed.

On the other hand for men, the presence and absence of landmarks did not induce any difference in both escape time and path distance, suggesting that men use geometric cues (Grön et al 2000).

Figure 2.2 shows the examples of the recorded path taken in the maze test at first time (0th week) men follow the more efficient path than women toward the goal in the maze without landmarks, but in the presence of landmarks this gender difference became smaller.

**Figure 2.2.**
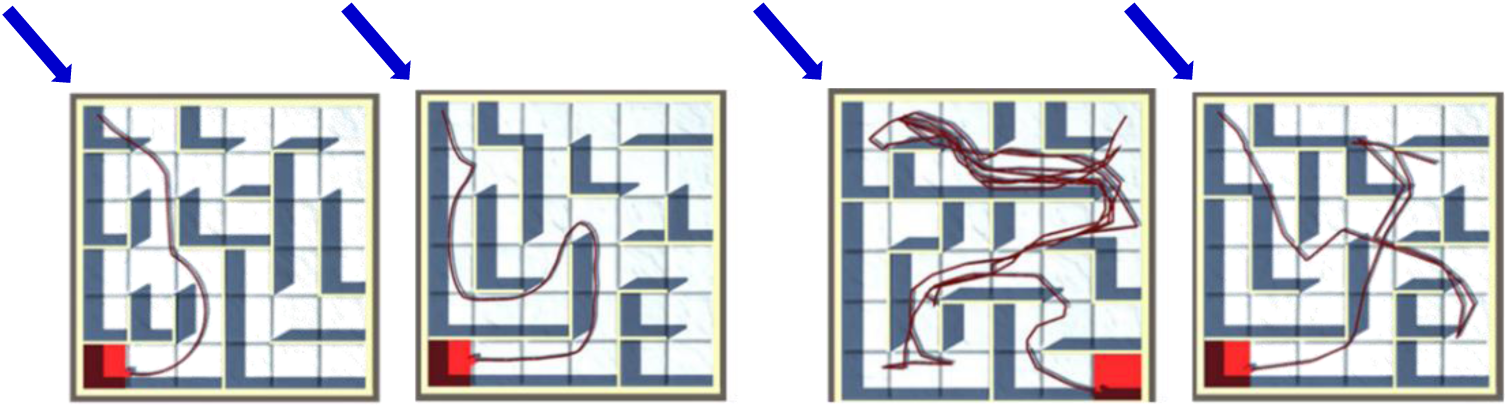
From left to right, young men without landmarks, young men with landmarks, young women without landmarks, young women with landmarks. Start (Arrow) and Goal (Red) are indicated.

## Section 3 Testosterone-Replacement Therapy (TRT) improved spatial cognition ability of LOH patients

In Section 2, the visuospatial navigation test successfully demonstrated differences in the spatial cognition ability between young men (with high T level) and young women (with low T level). The hippocampus is probably responsible to this spatial cognition ability from important previous reports. The spatial cognition ability of rats mainly depends on CA1 hippocampal neurons in which androgen receptor AR is localized (Tsien et al 1996) (McHugh et al 1996) (Tonegawa et al 1996) (Huerta et al 2000) (Tabori et al 2005) (Hatanaka et al 2015). Therefore, the observed difference in spatial cognition ability may significantly depend on the much higher hippocampal T levels in male than female as observed in rats (Hojo et al 2009) (Kato et al 2013). This hypothesis is dependent on the observation that nearly 20-fold T levels (in the hippocampus) and 100-fold T levels (in circulation) were observed in male rats than female rats (Hojo et al 2009) (Ooishi et al 2012) (Kato et al 2013).

It is shown also that T application to hippocampal CA1 glutamatergic neurons increased the number of synapses where memory is stored (Hatanaka et al 2015) (Murakami et al 2018) (Hojo & Kawato 2018). It may be therefore possible to find differences in maze navigation ability between individual men with high T and low T levels.

We applied spatial maze navigation task (with three landmarks) to LOH patients (with low T levels) in order to analyze a possible improvement in the spatial cognition ability upon TRT. T supplementation was performed by intramuscular injection of testosterone enanthate (TE, 250 mg) to LOH patients every 2 weeks, for 3 times.

### Materials and Methods

#### Subjects : LOH patients

This study was applied to men with symptoms of LOH who visited Male Menopause Outpatient Clinic of Teikyo University Hospital. They had at least one symptom of LOH including lethargy, general fatigue, malaise, depression, insomnia, sweating, hot flashes, dizziness, night sweat, sexual dysfunction, and decreased libido.

We assessed symptom scores with several specific questionnaires including the Heineman’s Aging males’ symptoms scale (AMS) for LOH, the international index of erectile function (IIEF-5) and self-rating depression scale (SDS) (Heinemann et al 2003) (Lunenfeld et al 2013) (Ide et al 2023). Endocrinologic data, including total T and free T were evaluated. All blood samples were collected between 09:00 and 11:00 to monitor endocrinological variables.

LOH was categorized by the AMS score (above **∼**45), total T levels (< ∼300 ng/dL) and free T levels (< 8.5 ∼ 11.8 pg/mL) (Lunenfeld et al 2013, Wu et al 2010) (Tsuru et al 2023) (Ide et al 2023).

Thirteen males diagnosed with LOH participated in the study. The average age was 52.3 ± 5.8 years old). Therefore, they were not older men (60-80 years), but rather middle-aged men.

The clinical study was approved by the Teikyo University Ethics Committee (Approval No. Teirin 15-148).

#### Serum hormone assay

Serum total T and luteinizing hormone (LH) were determined in the SRL Inc. (Tokyo, Japan) by Cobas 8000 electrochemiluminescence immunoassay (Hitachi High-Tech Co. Ltd., Tokyo, Japan), using ECLusys Testosterone Kit (Roche Diagnostics, Indianapolis, IN,USA). Serum free T was determined in the SRL Inc. by RIA, using the DPC Free Testosterone Kit (Mitsubishi Kagaku Iatron Inc., Tokyo, Japan) (Iwamoto et al 2009).

#### Procedures of TRT and maze navigation test

The task is conducted according to the following procedures (see Fig. 2.1).

1. After explaining the outline of the TRT for LOH, the subjects complete the AMS, SDS and IIEF5.
2. Show a video demonstration of the maze test (first time only).
3. Conduct the maze navigation test with three landmarks. The maze with three landmarks are used. This is because the maze with three landmarks is easier to try for LOH patients whose spatial cognition ability is lower than young men. When performing the maze test, maze number 1 should be entered for the first time, maze number 2 for the second time, and maze number 3 for the third time, to avoid mazes with identical goal positions.
4. The subjects are given 250 mg of testosterone enanthate (ENARMON DEPOT, ASKA Pharmaceutical Co. Ltd., Tokyo, Japan) by intramuscular injection for 3 times.
5. After a 2-week interval, repeat steps 1, 3 and 4.

The above procedure is repeated four times for each patient. The overall flow is shown in Figure 3.1.

**Figure 3.1.**
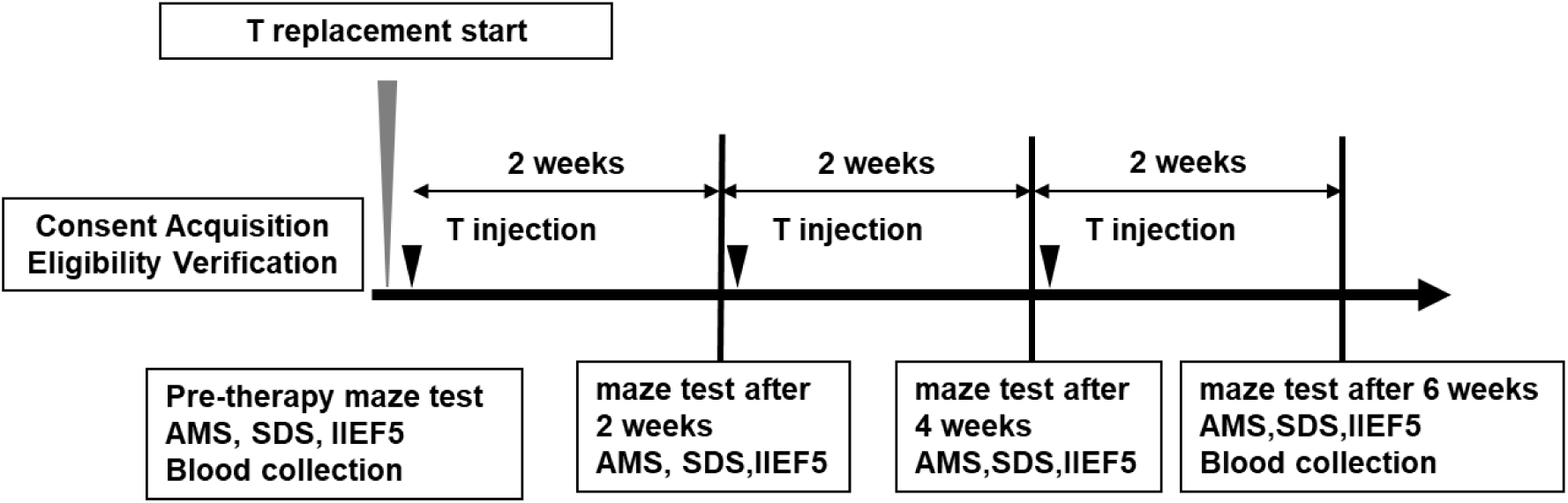
The overall flow of TRT and maze navigation test T is supplemented by intramuscular injection three times every two weeks. Blood samples are collected on the first and forth (final) maze test day in order to measure total T, free T, luteinizing hormone (LH), Hb and hematocrit (Ht) levels.

### Statistical Analysis

Data are expressed as mean ± SEM.

For analysis of navigation maze tests, we used a paired t-test. For analysis of AMS, SDS and IIEF-5 we used a paired t-test. For analysis of serum hormone levels, we used Wilcoxon signed rank sum test. A difference was considered significant at a value of ∗p < 0.05, ∗∗p < 0.001.

### Results

#### Improvement of maze navigation task scores by TRT

After 6 weeks TRT, with three times T injection every two weeks, both the maze escape time and the path distance are significantly shortened (Figure 3.2 and Figure 3.3).

**Figure 3.2.**
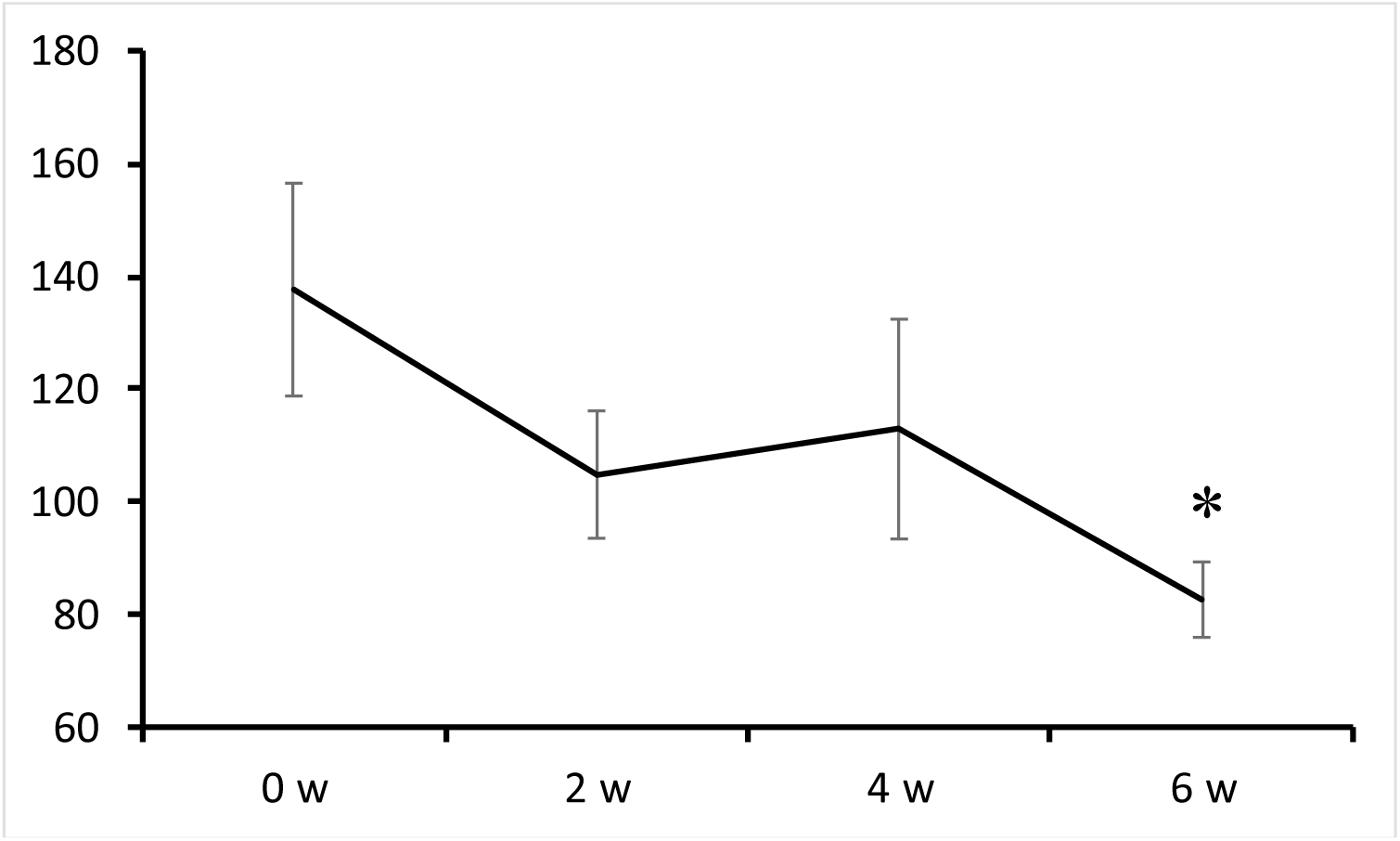
Biweekly maze test scores of escape time along TRT for 6 weeks. LOH patients (n = 13). Vertical axis is escape time (second).

**Figure 3.3.**
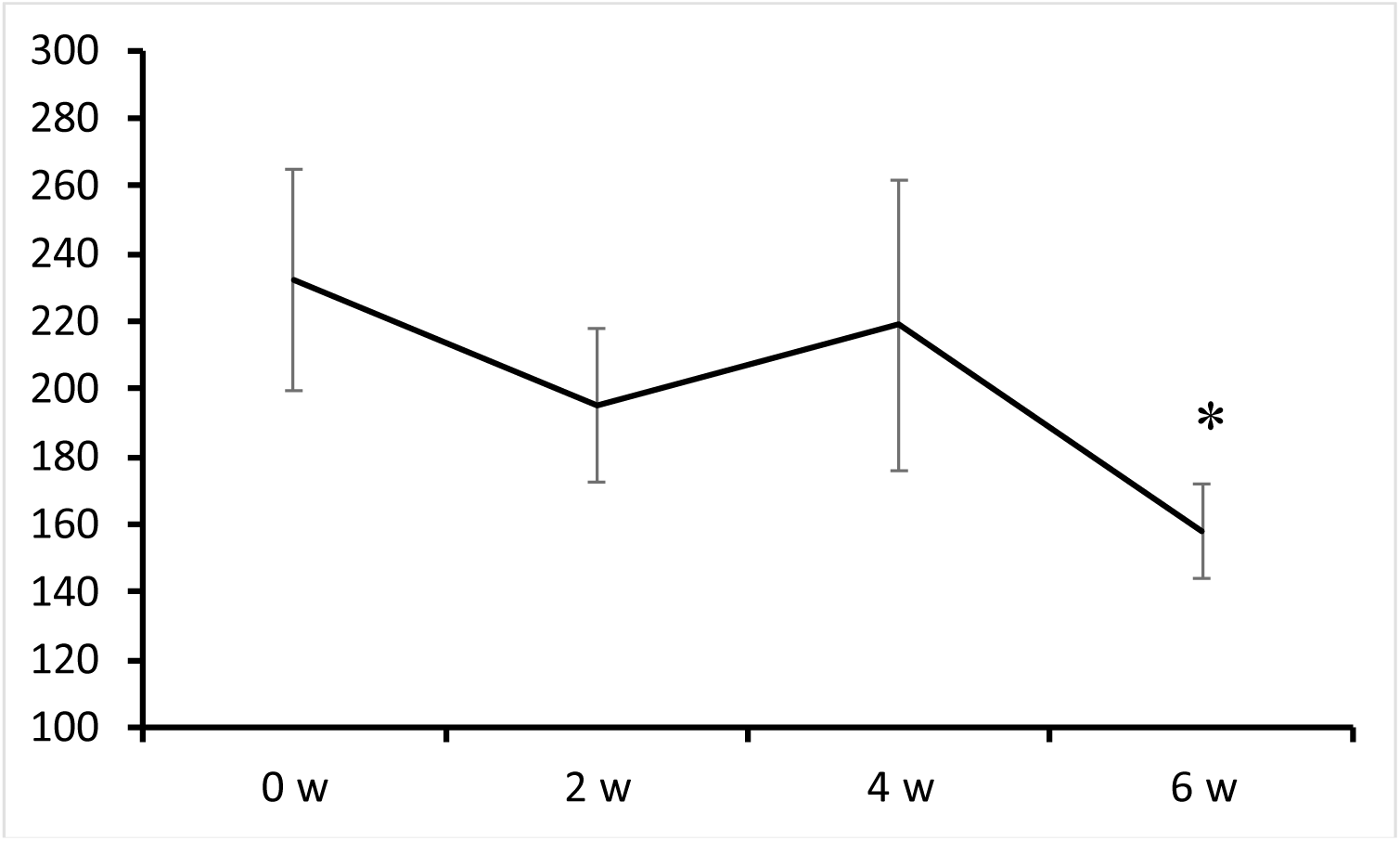
Biweekly maze test scores of path distance along TRT for 6 weeks. LOH patients (n = 13). Vertical axis is path distance (arbitrary unit).

A paired t-test revealed significant differences in the maze test scores before (0 week) and after 6 week treatments for both escape time and path distance.

Representative recorded paths of a typical LOH patient along TRT is shown in Figure 3.4.

**Figure 3.4.**
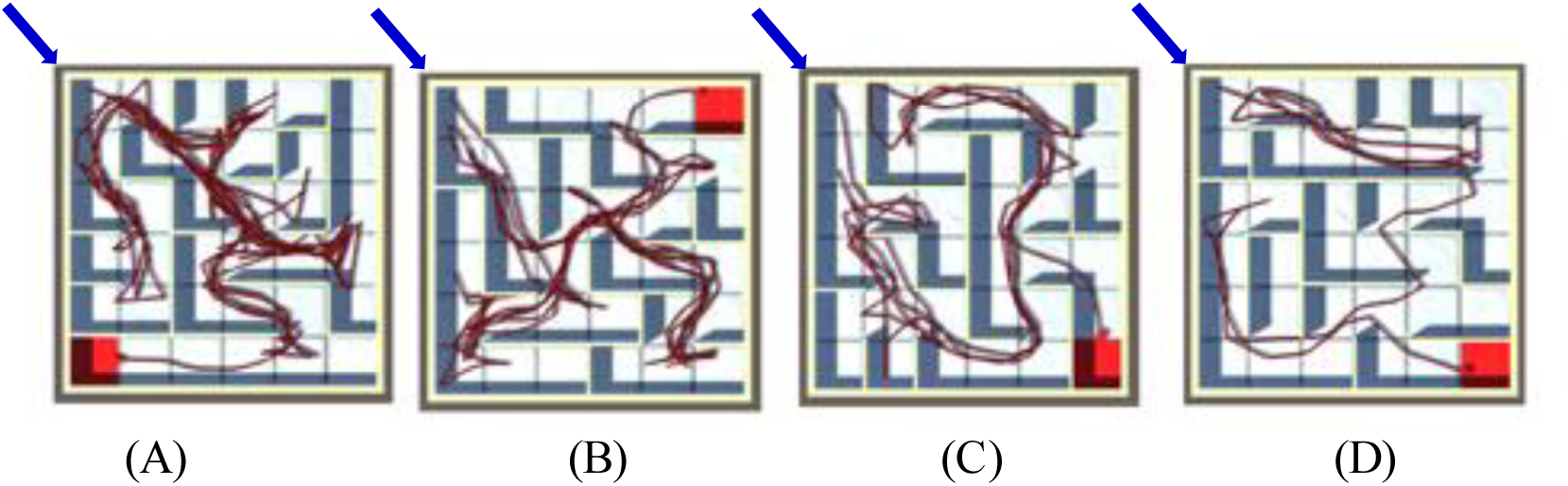
Recorded paths on the maze test obtained from one LOH patient. (A) before T-replacement (0 week), (B) after T-replacement (2nd week), (C) after T-replacement (4th week), (D) after T-replacement (6th week). Start (Arrow) and Goal (Red) are indicated.

#### Improvements of AMS, SDS and IIEF-5 scores after TRT

AMS showed a significant improvement, indicating the restorative effects of Aging males’ symptoms after TRT (Figure 3.5).

**Figure 3.5.**
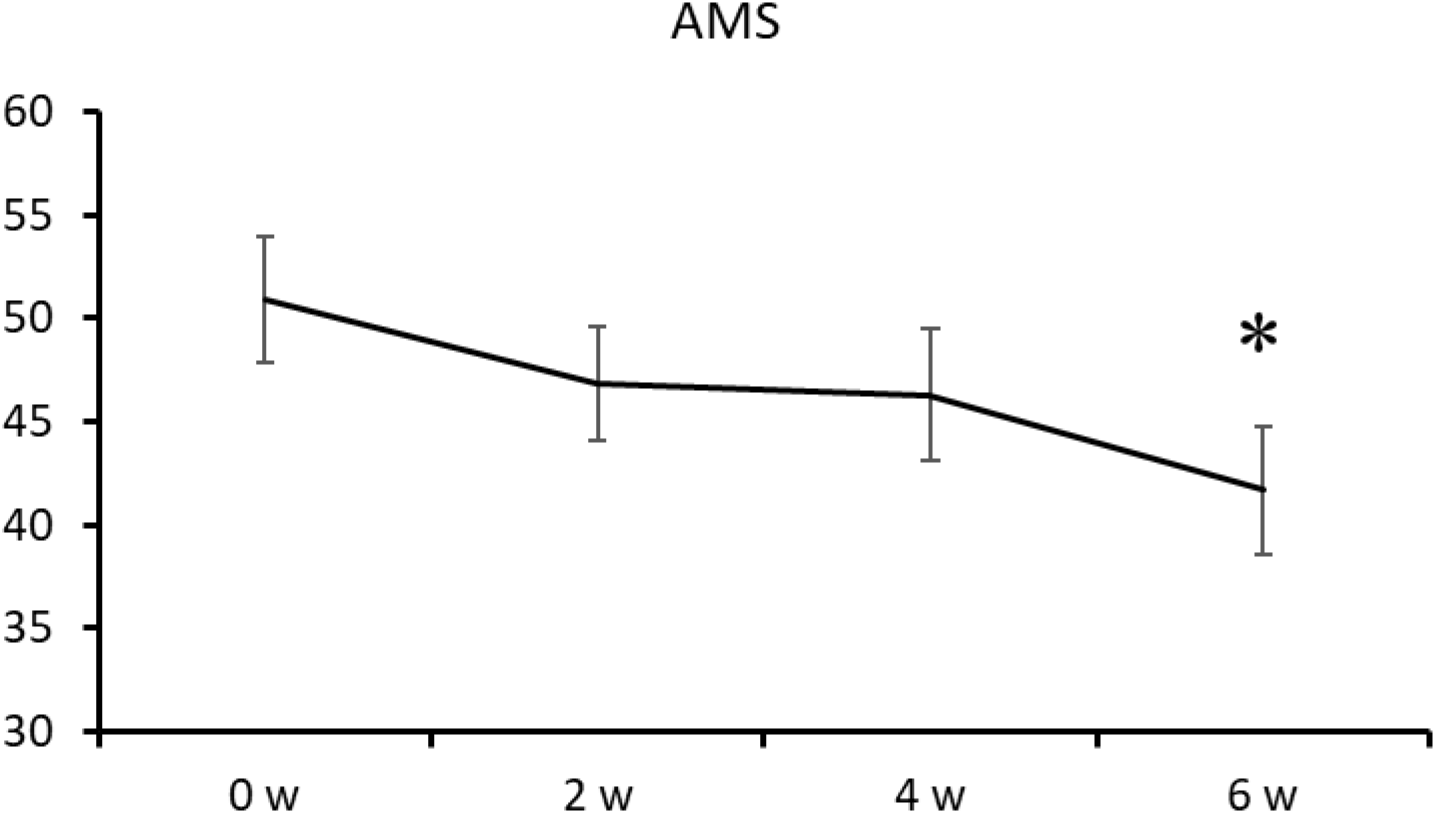
Biweekly scores of AMS along TRT for 6 weeks.

Other important markers, SDS and IIEF5, also showed improvement after TRT (Figures 3.6 and 3.7), indicating the recovery of depression and erectile function.

**Figure 3.6.**
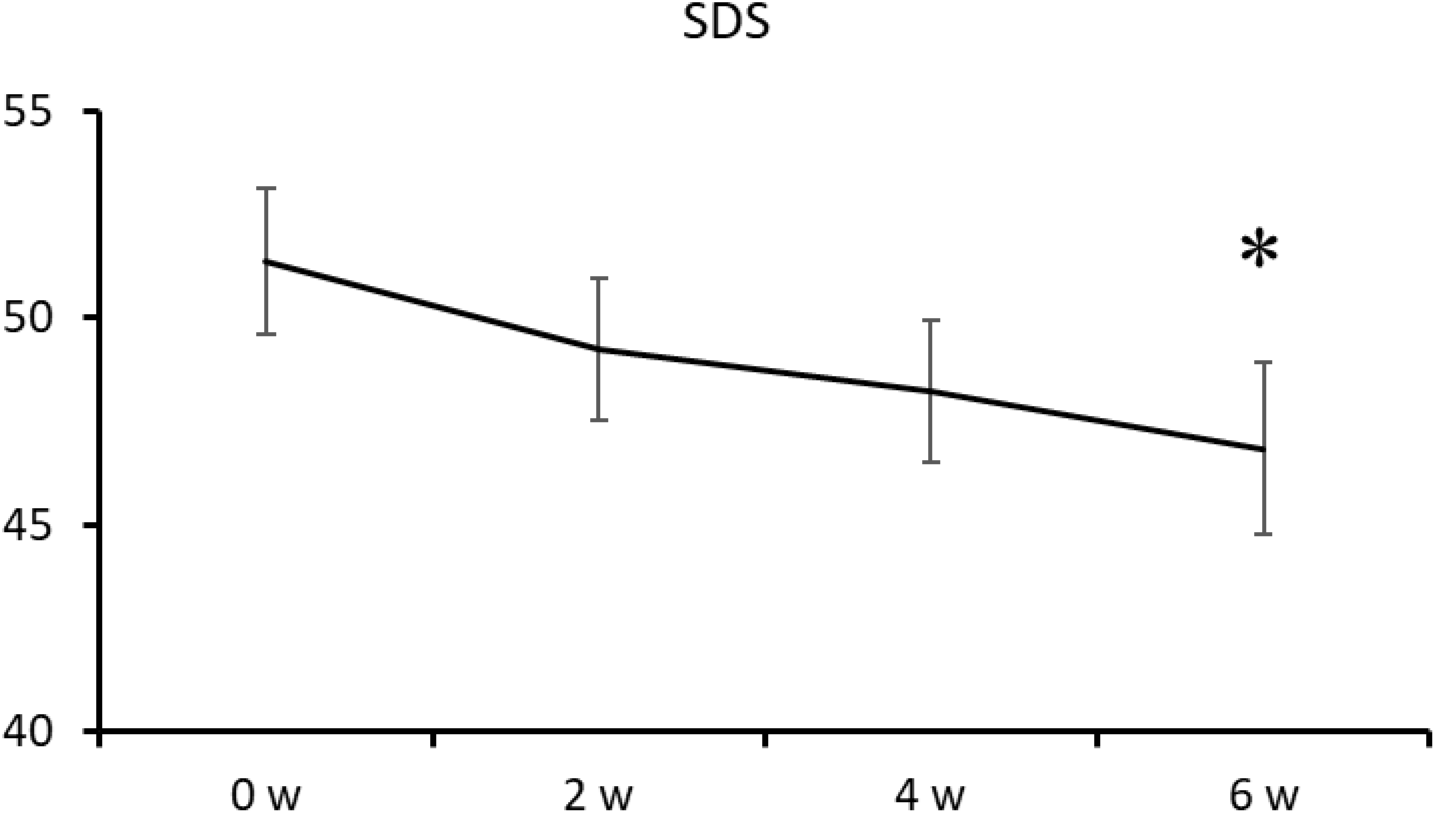
Biweekly scores of SDS (self-rating depression scale) along TRT for 6 weeks.

**Figure 3.7.**
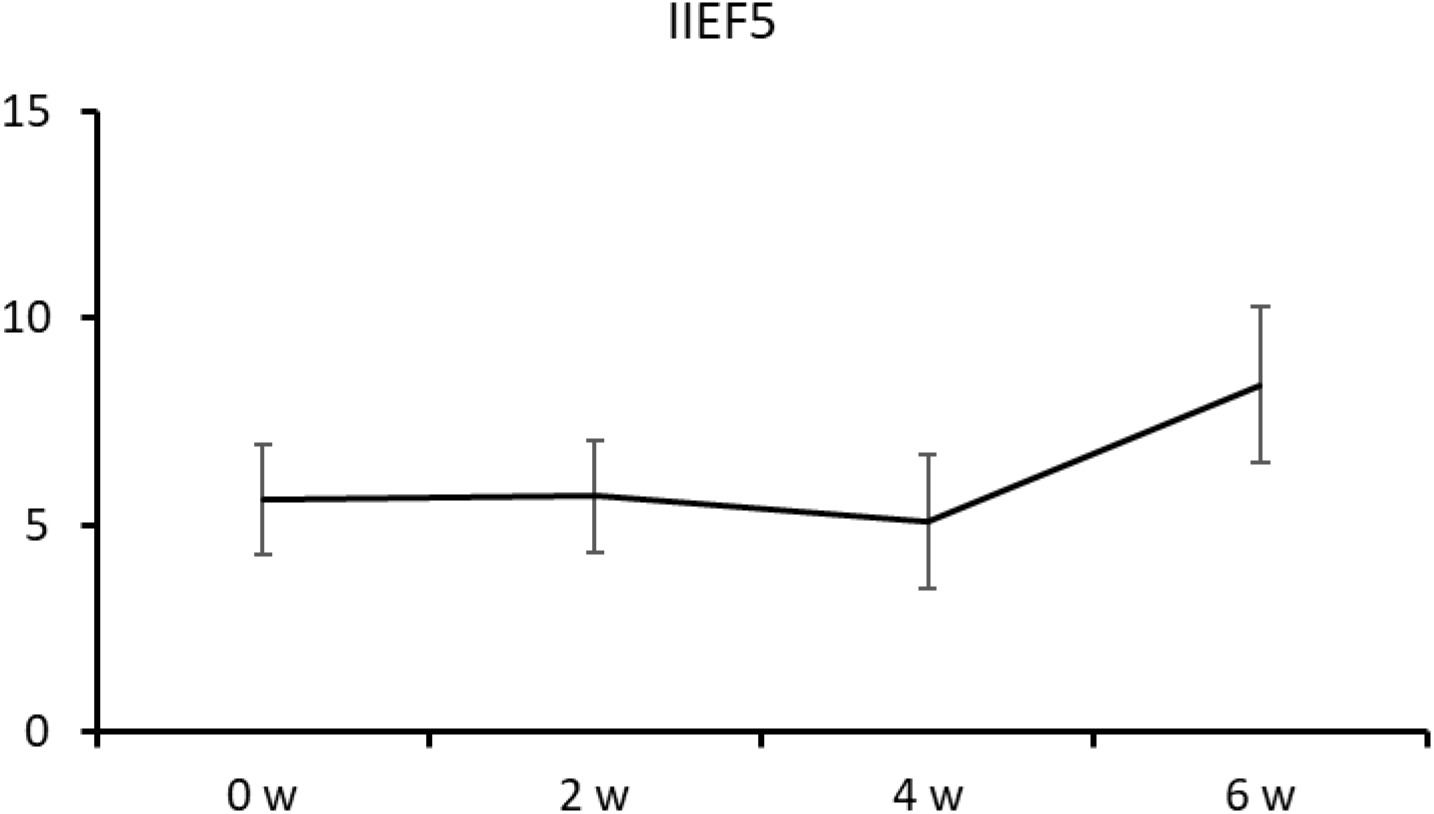
Biweekly scores of IIEF-5 (index of erectile function) along TRT for 6 weeks.

#### Changes in serum hormone levels

The results of blood tests before and after TRT indicated the significant elevation of total T and/or free T levels (Table 2).

**Table 2.** List of serum hormone levels and some other factors. Luteinizing hormone (LH), total T, free T, Hemoglobin (Hb) and hematocrit (Ht).

|  | Before TRT | 6 weeks after TRT | P value |
| --- | --- | --- | --- |
| LH (mIU/ml) | 4.8±0.6 | <0.1 |  |
| Total T (ng/dL) | 329.0±48.2 | 616.0±46.3 | P<0.01 |
| Free T (pg/ml) | 6.4±0.7 | 13.7±1.2 | P<0.01 |
| Hb (g/dL) | 14.5±0.2 | 15.3±0.4 | P<0.01 |
| Ht (%) | 43.1±0.7 | 46.3±1.0 | P<0.01 |

### Discussion

In the present study, spatial navigation ability was shown to be improved by elevation of the T levels in LOH patients by TRT. This might be caused by increase in the T levels in the brain hippocampus including CA1 neurons in which AR is localized and which are responsible to spatial cognition (Tsien et al 1996) (Tonegawa et al 1996) (Tabori et al 2005) (Hatanaka et al 2015).

The present results might be consistent to the previous observations that (1) spatial navigation ability (that is hippocampus-dependent function) is one of the few cognitive functions for which male performance is superior to female performance (Grön et al 2000), and that (2) the much higher T levels in male hippocampus than that in female hippocampus (e.g., approximately 100-fold higher) could support the better performance of male than female subjects (Tsien et al 1996) (Tonegawa et al 1996) (Hojo et al 2009) (Kato et al 2013) (Hatanaka et al 2015) (Kuwahara et al 2021) (Spritzer et al 2021).

Since LOH patients have lower T levels than those in healthy men, the present TRT-induced T elevation in LOH patients probably improved the spatial navigation task scores (e.g., escape time and path length) (Figures 3.2 and 3.3).

So far, this kind of investigation, particularly focusing on spatial navigation task for LOH patients, has not been performed intensively. Besides spatial navigation, there are other several cognitive abilities, e.g. attention, memory, calculation, verbal ability, auditory and visual processing. However, many previous studies suggest that these general cognitive abilities did not show significant improvement in older men (60 – 80 years old) by long term TRT (for half a year or two years) (Borst et al 2014) (Lisco et al 2020) (Gregori et al 2021). Importantly, the LOH patients in the present study are not older men but rather middle-aged men (between 45 and 60 years old) who have more neural plasticity than older men, and this may be one reason why the present short term TRT (6 weeks) showed effective improvements.

LOH patients exhibit symptoms that include hot flashes, depression, fatigue, and decreased libido due to decreased total T and/or free T. The main symptoms of LOH are well evaluated by the AMS, SDS, and IIE5 scores. We observed significant improvement of AMS, SDS, and IIE5 scores by TRT (Figures 3.5, 3.6), which correlated well with the improvements of spatial navigation scores (escaped time and path length) (Figures 3.2, 3.3).

Importantly, in Section 3, subjects (LOH patients, middle-aged) did not show mild cognitive impairment (MCI) symptoms or dementia. MCI symptoms were examined using the Revised Hasegawa’s Dementia Scale (HDS-R) that is a simplified Mini-Mental State Examination (MMSE) (Gong et al 2021) (Arevalo-Rodriguez et al 2015). LOH subjects in Section 3 showed no MCI with almost full score in HDS-R, and this is probably because subjects were younger than 61 years old.

Interestingly, many studies recently reported that the long term TRT (e.g., for 2 years) for older patients (aged 65 years or older) did not significantly improve their cognitive impairment (as judged from MMSE or HDS-R test), though TRT showed significant improvement in muscle strength and physical performance (Lisco et al 2020).

In the first part of this study, it was important to select and create the suitable spatial cognition test in order to examine the TRT effect on male LOH patients.

Many researches were conducted on the spatial cognitive functions of men and women (Hyde 2016), and it is generally accepted that men outperform women. For example, men show higher scores than women in spatial route-learning task, virtual maze task, and visuo-spatial task (Galea & Kimura 1993) (Moffat et al 1998) (Bosco et al 2004).

On the other hand, there are also other types of spatial cognition tests including the Mental Rotation Test (MRT) (Feng et al 2007) (Neubauer et al 2010). In MRT, subjects should determine which objects are identical by rotating several objects in their mind.

By using MRT, men showed superior results to women in spatial cognition. However, little is known about specific brain regions and gender dependent regions which are excited during MRT. Therefor we cannot further analyze deeply T-dependence of MRT.

Therefore in the present study, we chose spatial route-learning task which is suitable to evaluate men’s cognitive ability that depends on T levels. Fortunately, spatial navigation solely depends on the hippocampal CA1 neural function and AR activity which are closely correlated with T levels (Tsien et al 1996) (Tonegawa et al 1996) (Hojo et al 2009) (Kato et al 2013) (Hatanaka et al 2015) (Spritzer et al 2021).

## Data Availability

All data produced in the present work are contained in the manuscript.

## Acknowledgement

Department of Cognitive Neuroscience, Teikyo Univ. is an endowment department, supported with an unrestricted grant from Japan Tobacco Inc.

## Author Contributions

SK, HM, MK and SH conceived and designed the study. HM, YK, MK, MO-I and M Soma conducted the experiments and analysis of the data. SK, HM and YK wrote the manuscript. SH, MO-I and M Saito contributed to discussions. All authors provided feedback on the manuscript.

